# Using Digital Humanities for Understanding COVID-19: Lessons from Digital History about earlier Coronavirus Pandemic

**DOI:** 10.1101/2022.02.02.22270333

**Authors:** Tado Jurić

**Affiliations:** Catholic University of Croatia

**Keywords:** Google Ngram, Big Data, epidemic, COVID-19, Russian flu, Digital Humanities

## Abstract

**Background:** At the time of the COVID-19 epidemic, it is useful to look at what lessons (digital) history can give us about the past pandemics and dealing with them. We show that the Google Ngram (GNV) can discover hidden patterns in history and, therefore, can be used as a window into history. By using the approach of Digital Humanities, we analysed the epidemiological literature on the development of the Russian flu pandemic for hints on how the COVID-19 might develop in the following years.

**Objective:** Our study is searching for evidence that the COVID-19 is not a unique phenomenon in human history. We are testing the hypothesis that the flu-like illness that caused loss of taste and smell in the late 19th century (Russian flu) was caused by a coronavirus. We are aware that it is difficult to formulate a hypothesis for a microbiological aetiology of a pandemic that occurred 133 years ago. But differentiating an influenza virus infection from a COVID-19 patient purely on the clinical ground is difficult for a physician because the symptoms overlap. The most crucial observation of similarities between the Russian flu pandemic and COVID-19 is the loss of smell and taste (anosmia and ageusia). The objective was to calculate the ratio of increasing to decreasing trends in the changes in frequencies of the selected words representing symptoms of the Russian flu and COVID-19.

**Methods:** The primary methodological concept of our approach is to analyse the ratio of increasing to decreasing trends in the changes in frequencies of the selected words representing symptoms of the Russian flu and COVID-19 with the Google NGram analytical tool. Initially, keywords were chosen that are specific and common for the Russian flu and COVID-19. We show the graphic display on the Y-axis what percentage of words in the selected corpus of books (collective memory) over the years (X-axis) make up the word. To standardise the data, we requested the data from 1800 to 2019 in English, German and Russian (to 2012) book corpora and focused on the ten years before, during and after the outbreak of the Russian flu. We compared this frequency index with “non-epidemic periods” to test the model’s analytical potential and prove the signification of the results.

**Results:** The COVID-19 is not a unique phenomenon because the Russian flu was probably the coronavirus infection. Results show that all the three analysed book corpora (including newspapers and magazines) show the increase in the mention of the symptoms “loss of smell” and “loss of taste” during the Russian flu (1889-1891), which are today undoubtedly proven to be key symptoms of COVID-19.

In the English corpus, the frequency rose from 0.0000040433 % in 1880 to 0.0000047123 % in 1889. The frequency fell sharply after the pandemic stopped in 1900 (0.0000033861%). In the Russian corpus, the frequency rises from 0 % in 1880 to 0.0000004682 % in 1889 and decreased rapidly after the pandemic (1900 = 0.0000011834 %). In the German corpus, the frequency rose from 0.0000014463 % in 1880 to 0.0000018015 % in 1889 and decreased also rapidly after the pandemic (1900 = 0.0000016600 %).

According to our analysis of historical records with the approach of GNV, 1) the ‘natural’ length of a pandemic is two to five years; 2) the pandemic stops on their own; 3) the viruses weaken over time; 4) the so-called “herd immunity” is not necessary to stop the pandemic; 5) history has shown that a significant crisis does not need to occur after the COVID-19 pandemic.

**Conclusion:** According to our study, the Google Books Ngram Viewer (GNV) gives a clear evidence of the influence that social changes have on word frequency. The results of this study open a discussion on the usefulness of the Google Ngram insights possibilities into past socio-cultural development, i.e. epidemics and pandemics that can serve as lessons for today. We showed hidden patterns of conceptual trends in history and their relationships with current development in the case of the pandemic COVID-19.

The benefit of this method could help complement historical medical records, which are often woefully incomplete. However, this method comes with severe limitations and can be useful only under cautious handling and testing. Despite the numerous indications we have shown, we are aware that this thesis still cannot be confirmed and that it is necessary to require further historical and medical research.

## Introduction

An answer to the question of the future development of the COVID-19 pandemic is of high importance for all societies and countries worldwide. By messages from the media and official reports, we know so far that they are unreliable and that epidemiological predictions are uncertain. Because medical evidence and epidemiological estimates can not answer this question, looking at history’s lessons is helpful.

Studying past pandemics shows that elements relevant to the COVID-19 pandemic are repeated, and that the measures that we undertake today are precisely the same as what they did in Spanish flu and partially in Russian flu – social distancing, wearing masks, quarantining, travel restrictions [1].

But just as individuals forget the past, so do societies [2]. This paper shows that Digital Humanities approaches might be used to track historical epidemics and renew knowledge from the past.

According to Brüssow^3^ and Van Ranst [1], the Russian flu might have been a coronavirus infection. Due to the limitations, it is impossible to have medical evidence for this thesis. Therefore, we set the hypothesis that the tools of Digital Humanities, especially Google Books Ngram Viewer (GNV), can be helpful to find the clinical data from the historical reports.

Our goal in this paper is to analyse the epidemiological literature on the development of the Russian flu pandemic (1889) for hints on how the COVID-19 might develop in the following years and compare the similarities. The historical record of past pandemics might thus provide us with the so-called “retrodictions” [3] on possible future scenarios for the COVID-19 pandemic.

According to Van Ranst, the first coronavirus was transmitted from bovine to humans. According to this thesis, what we are experiencing today has already been experienced in the late 19th century [1]. To find evidence, we have analysed the indices by the clinical data from the historical reports from the Google corpus of digitised books that includes 15 million books (12% of all books ever published). We asked ourselves especially if the COVID-19 pandemic is a unique occurence in humanity, whether it will disappear or become endemic and what the future consequences will be like.

According to our study, the GNV clearly shows the influence that social changes have on word frequency. The relationship between values fostered in a society and its language is close.^4^ Our basic assumption is, that when culture and language are linked, then one should impact the other. Furthermore, it has been recently shown that during seasonal influenza epidemics users of Google are more likely to engage in influenza-related searches and that this signature of influenza epidemics corresponds well with the results of CDC surveillance [5]. We, therefore, reasoned that the Big Data and Digital Humanities approaches might be used to track historical epidemics and give us answers to some questions that would otherwise remain unanswered.

### Big Data in the Digital Humanities

The expression “Big Data” has been spreading since 2011. The term is used in academia, industry and the media, but it is not even today exactly clear what it means. Is it an object of study, a method, a group of technologies or a discipline?^6^ In general, the definitions combine two essential ideas: storage of a large volume of data and analysing this data quantitatively and visually to find patterns, establish laws and predict conduct [7]. The classic definition of “Big Data” is a formula - the three “Vs”: Volume, Velocity (data that is constantly generated) and Variety (texts, images, sounds) [7].

According to Oza, Digital Humanities is “a broad field of research and scholarly activity covering the use of digital methods by arts and humanities researchers and how the arts and humanities offer distinctive insights into the major social and cultural issues raised by the development of digital technologies” [8]. Work in this field is methodological and interdisciplinary in scope, involving multiple skills, disciplines, and areas of expertise with the investigation, analysis, synthesis and presentation of data in electronic form [8]. According to Burdick et al., “Digital Humanities is less a unified field than an array of convergent practises that explore a universe in which print is no longer the primary medium in which knowledge is produced and disseminated [9].

Big Data is widely used today in digital culture as a promising method for deriving new understanding from massive aggregations of information. The ability to collect a vast amount of data from text, images, and media and to analyse it using computerised algorithms creates endless opportunities in many areas [10]. “Big data” methodologies bring new potential not just for medicine and business analytics but also for humanities research and social sciences. Latour believes that big data can resolve the gap between the micro and the macro in sociology, the unexplained relations between macro-social phenomena and the individuals taking part in that phenomena [11].

In the humanities, one can only speak of Big Data in connection with the technologies associated with this phenomenon, such as data mining, stylometry or natural language processing [12]. It is crucial to differentiate between “data”, “raw material”, and “information”. According to Castro, more than the finished product, what matters in the Digital Humanities is the creative process when a phenomenon is “modelled”. The aim is to gain new knowledge and meanings by generating an external object that represents it [12]. Humanistic disciplines such as history, philosophy, and philology are characterised by a specific object of study and a method that seeks to understand particular, unusual and even unique cases through text commentary. According to Castro, Big Data in humanities will unquestionably affect certain clichés about the humanities and their classic objects of study [12]. Although the tool may help develop specific theories concerning socio-cultural phenomena, many researchers claim that the data obtained with Google Books Ngram Viewer is not reliable enough to confirm these theories (see restrictions) [13].

### Google NGram

Reading small collections of carefully chosen works enables scholars to make robust inferences about trends in human thought. However, this approach rarely allows precise measurement of the underlying phenomena. According to Michael et al. [14], computational analysis of the digitalised corpus of books enables us to observe cultural trends and subject them to quantitative investigation. This new field, *Culturomics*, extends the boundaries of scientific inquiry to a wide array of new phenomena [15].

One of the tools that serves Digital Humanities is GNV [16]. This tool has been created on top of Google Books, the largest digitised collection of books. GNV is creating a graphical representation of the frequency of occurrence of search terms over the years in a selected corpus of digitised books [14]. It contains a corpus of over 15 million digitised books and over 600 billion words in 2022. It is actually the world’s largest archive - which is also available online and for free. Google states that its team, together with Cultural Observatory, Harvard University, Encyclopaedia Britannica and the American Heritage Dictionary, have digitised over 15 % of all books that have ever been published from over 40 university libraries (such as the University of Michigan and the New York Public Library) and individual publishers [16]. In 2004, Google began with scanning books (OCR). The first version in 2009 had six million books; in 2012, the second version incorporated eight million books [17], and the 2019 version had over 15 million books. Due to the wide scale of digitally archived texts, these corpora are not limited to specific genres. It includes all sorts of literature, ranging from academic publications to biographies and novels [18]. The collection contains books dating back to as early as 1473 and texts in 478 languages [19]. Of the 15 million books scanned, the country of publication is known for 91.5%, authors for 92.1%, publication dates for 95.1%, and the language for 98.6%. The OCR quality is generally higher for the languages that use a Latin alphabet (English, French, Spanish, and German), and more books are available [19].

The new version of GNV from 2019 is characterised by improved optical character recognition (OCR) and better underlying library and publisher metadata [20]. Google estimates that over 98% of words are correctly digitised for modern English books [14]. The GNV does offer differentiation by language. Subcorpora exist for eight languages, with the English corpus being the biggest, containing more than 350 billion words. The corpus covers a period from 1500 until 2008. However, Michel et al. point out that search inquiries between 1800 and 2000 will deliver the highest data density and quality [19]. The problem is that smaller language communities are not included.

Compared to other big data sets, the Ngram Viewer enables a fast and easy access to this pool of information [18]. Next to a regular search field for the term or phrase of interest, the online tool offers filtering options for the period, the language, the degree of smoothing that affects how the graphs of the search result are displayed, and a case insensitive option. It is also possible to search for more than one term or phrase for direct comparison [18]. To avoid overwhelming the diagram in any given year, the graph will only show books with the term(s) if there are more than 40 occurrences. To deal with the problem presented by the increase in published books over time, the results are normalised by the number published each year [21].

Without a normalisation, it would be impossible to compare the frequency of a specific ngram over time, as the number of books published in 1500 is not equal to the number of books published in 2010 [21]. The viewer, therefore, displays a percentage of the number of occurrences, where the percentage is calculated out of the total number of books published in a given year. Clicking on a point in the plotted graph shows the percentage of occurrences for that year [10]. The data generated by specific inquiries can then be exported as a list and processed with alternative software packages (for example, “R”), particularly with spreadsheet applications [18].

GNV can be used as a tool for discovering hidden patterns of conceptual trends, trends in knowledge, the relative importance of concepts etc. [22]. The main challenge for Digital Humanities will be to take patterns discovered by digital analysis and discern correlations to historical events, to explain patterns by historical forces, causes and relations [10].

### How to use NGram in Digital Humanities

The GNV calculates how often a certain n-gram appears in the selected corpus of a given year relative to the total number of n-grams [14]. In computational linguistics, an Ngram is a contiguous sequence of *n* items from a given sequence of text [16]. The items can be phonemes, syllables, letters or words. The GNV database supports n-gram sequences of up to five elements [10].

For example, “I” is a 1-gram and “I am” is a 2-grams. This means that if the researcher searches for one word (unigram), he will get the percentage of this word to all the other words found in the corpus of books for a specific year [22]. If the researcher entered more than one word or phrase, each one is represented by a colour-coded line to contrast with the other search terms. This is similar to Google Trends [23], with the exception that the search covers a longer period [24].

The researcher can modify searches by time frame, degree of detail and corpus type, including several different languages as mentioned. As well as verbs and nouns, scholars can also search for adjectives, adverbs, pronouns, determiners, prepositions and more, using the tags listed on this helpful page of tips. Google estimates the accuracy of this tagging at 95% [22].

A few features of the GNV may appeal to users who want to dig a little deeper into phrase usage: *wildcard search, inflexion search, case insensitive search, part-of-speech tags* and *ngram compositions*. [16] For comparisons of several n-grams, it is possible to combine or separate two expressions and divide or multiply expressions to compare n-grams of different frequencies or to isolate frequencies of one n-gram in relation to another. Adding a “+” operator between n-grams allows the researcher to combine multiple frequencies into one. Adding the operator “-” between n-grams allows the subtraction of frequencies from the right from the frequencies from the left and thus enables the measurement of frequency connectivity [14].

Adding the “/” operator between n-grams allows isolating the movement of one frequency to another. Adding the operator “*” between n-grams multiplies the frequency on the left by the frequency with the selected value, that is, by the given number. It allows a comparison of two distinctly different frequencies. Adding the “:” operator between n-grams uses the n-gram on the left and the corpus on the right, and compares n-grams in different corpora [16].

Representation of words in multiple grammatical categories can be achieved by adding the code “_INF” as a suffix to the word’s root. Example: “book_INF” generates the appearance of words such as “books”, “booking”, “booked” for viewing in a single graphical display [16]. GNV offers the option to tag words in search such as “_NOUN_” (noun), “_VERB_” (verb), “_ADJ_” (adjective), “_ADV_” (adverb). These labels can serve as part of a word or make up the word itself. By entering the operator “=>,” it is possible to show the relationship between words and their connection in a sentence.

There is also a case-insensitive option - displaying words written in lowercase, uppercase only, or a combination of words [14]. If smoothing factor “1” is selected as the smoothing level, it means that the data are shown for - for example, 1990 will be the average of the raw data for 1990 summed with one value on each side (previous and future years) and divided by the number year (data for 1989 + data for 1990 + data for 1991) [25].

GNV does not make the search result available for further processing. Even though it is possible to download the raw data, this option only addresses extensive scale analyses that require technical resources and advanced know-how in computer science [26]. However, there is a pragmatic way of extracting data from the HTML source code shown by Chumtong and Kaldewey [18].

The basic method used by GNgram is text mining. It is a method for gathering structured information from unstructured text discovering meaningful relationships [27]. Text mining has significant potential for academic application [27] to 1) develop new hypotheses, 2) systematic reviewing of literature, and 3) testing of hypotheses. Documents can be mined to confirm or deny an existing hypothesis. In many cases, this might be the first opportunity to test an established belief about something [27].

Text mining enables the identification of patterns and relationships within a large body of texts that would otherwise be extremely difficult or time-consuming to discover. Therefore, it is a method that can speed up research and allow us to pose new questions or test the old ones.

One of the merits of this tool is that it enables the socio-cultural researcher to spend more time analysing data than on their collection, which is usually very time-consuming [13].

According to Zięba, since the lexical changes are gradual and relatively stable, the fluctuations in word frequency are relevant, and their study will improve our comprehension of the social changes and their consequences. [13] However, this method comes with severe limitations and can be useful only under the condition of cautious handling and testing. Otherwise, there is the high potential to gather garbled or false results due to badly formed questions being asked of data or due to the nature of the text(s) under study [27]. It is important to stress that no result from text mining should be taken at face value for historians. It is essential that results be checked and confirmed, and this often involves manually delving into the text under study (see limitations and methodology).

### Literature review

Since its introduction in 2010, GNV has been widely described and applied both in the social and natural sciences [13]. Berry (2012) describes it as an example of “the way in which code and software become the conditions of possibility for human knowledge” [27]. Rutten et al. treat it as a tool to overcome a “chronological distance, or time lag, between books and their subject matter in studies of memory” [28]. Michalski et al. (2012) suggest the GNV could be used “as a fast prototyping method for examining time-based properties over a rich sample of literary prose” [29].

Linguists used it to investigate biomedical domain literature in respect of terminology changes. In social studies, it was used to prove that moral ideals and virtues decreased significantly in the American public conversation, to analyse the concepts of happiness across time and cultures, to trace the roots of industrial ecology education to the 1960s and 1970s, to study the relations of science and capitalism, to trace the history of marketing and to introduce the concept of information overload [13].

As mentioned, Michel et al. [14] showed that the corpus enables investigators to study cultural trends quantitatively. The authors inquire into collective memory, compare the rise and fall of fame of the most well-known people, and uncover censorship in Nazi Germany [14]. Michel et al. showed that this approach could provide insights into fields as diverse as lexicography, the evolution of grammar, collective memory, the adoption of technology, the pursuit of fame, censorship, and historical epidemiology. The authors examined timelines for four diseases: influenza, cholera, HIV and poliomyelitis. In the case of flu, peaks in cultural interest showed excellent correspondence with known historical epidemics (the Russian Flu of 1890, the Spanish Flu of 1918 and the Asian Flu of 1957) [19].

Newberry et al., 2017 uses Google Ngram Viewer to analyse changes in the English language from the 12th to the 21st century [30]. Greenfield tested with GNV her theory on the influence of individualism on the individual’s values, behaviour, and psychology [31].

Acerbi et al. explored the presentation of emotions in books through the twentieth century using GNV. The authors conclude that stressful and violent historical events leave traces in the expression of emotions in books, so it is possible to detect “happy” and “sad” periods of history, depending on the representation and use of words for certain emotions through books [32]. Overall, GNV has allowed scholars to shed further light on various topics such as gender differences [33], emotions [34], personality [35], cognition [36], psychotherapy [37], moral values [38], education [39], nature [40], and the development of individualism and collectivism [41].

### Epidemics through history

Epidemics and pandemics have always been a part of human life. Since the existence of man, there have been infectious diseases. According to Harari, infectious diseases start when a person begins living sedentary; stops collecting and hunting. The First Agrarian Revolution cost man various diseases and contagions. The man no longer moved; he began to breed, keep animals and live in one place, which became an excellent prerequisite for developing diseases [42].

The spread of trade and the interaction of a growing number of people has led to epidemics, and in those times, it was not even known what humanity was facing. As humanity became more civilised, with the emergence of larger cities and population growth, exotic trade routes, and increased contact with different people, animals and ecosystems, the emergence of pandemics became greater [43].

The infographic below outlines some of the deadliest pandemics in human history, from the Antonine Plague that struck the Roman Empire from 165 to 180 to today’s current events and coronaviruses.

By the end of the 16th century, influenza was likely beginning to become understood as a specific, recognisable disease with epidemic and endemic forms. Since pandemic 1781–1782, starting in China, influenza became associated with sudden outbreaks of febrile illness [44].

Around the world, during the pandemics of 1889 (Russian flu) and 1918/1919 (Spanish flu), between 50 and 100 million people are estimated to have died. A direct comparison between the pre-pandemic and the coronavirus cannot be made. The world at the time did not know what made people die, and viruses as the cause of the disease were discovered only in 1933. But these pandemics still have something in common: they have thrown humanity into a deep crisis. That is why we wonder if the experiences from historical records about pandemics can help us prepare for the actual pandemic and the time after the pandemic.

The problem also arises in differentiation between Flu and COVID-19. Flu and COVID-19 are contagious respiratory illnesses, but different viruses cause them. COVID-19 is caused by infection with a coronavirus first identified in 2019, and flu is caused by infection with influenza viruses [45]. Similarities are that both COVID-19 and flu can have varying signs and symptoms, ranging from no symptoms (asymptomatic) to severe symptoms. Common symptoms that COVID-19 and flu share include: fever or feeling feverish/having chills; cough; shortness of breath or difficulty breathing; fatigue (tiredness); sore throat; runny or stuffy nose; muscle pain or body aches; headache; vomiting and diarrhoea [45].

### Russian flu - an earlier coronavirus pandemic

According to Van Ranst, flu-like illness that caused loss of taste and smell in the late 19th century was probably caused by a coronavirus that still causes the “common cold” in people today [1]. According to Van Ranst, COVID-19 virus will follow a similar pattern and become a continuously circulating, or ‘endemic’ virus, joining four other human coronaviruses that infect people with common cold symptoms. “The virus OC43 is still around. It is now responsible for common colds (…). And probably in some elderly people, it can lead to severe illnesses (…). COVID-19 is now the most intensely studied virus ever. These other viruses received far less attention” [1].

Vijgen et al. [46] showed that at the same time historical records showed a highly infectious respiratory disease with a high mortality rate affecting cattle herds around the world [47]. Today, the same similar disease is known as contagious bovine pleuropneumonia [46]. In the XIX century, the clinical symptoms of CBPP would have been difficult to distinguish from those of BCoV pneumonia. Most industrialised countries mounted massive culling operations between 1870 and 1890 and were able to eradicate the disease by the beginning of the XX century [48]. According to Vijgen et al., during the slaughtering of CBPP-affected herds, there was ample opportunity for the culling personnel to come into contact with bovine respiratory secretions [46]. Around the period in which the BCoV interspecies transmission would probably have taken place, a human epidemic ascribed to influenza was spreading worldwide.

The 1889–1890 pandemic probably originated in Central Asia [49] and was characterised by malaise, fever, and pronounced central nervous system symptoms [50]. Absolute evidence that an influenza virus was the causative agent of this epidemic was never obtained due to the lack of tissue samples from that period [46]. However, post epidemic analysis in 1957 of the influenza antibody pattern in sera of 50 to 100 years old indicated that H2N2 influenza antibodies might have originated from the 1889–1890 pandemic.^51^ According to Vijgen et al., dating the most recent common ancestor of BCoV and HCoV-OC43 to around 1890 is one argument. Another argument is that central nervous system symptoms were more pronounced during the 1889–1890 epidemic than in other influenza outbreaks [46]. It has been shown that HCoV-OC43 can be neuroinvasive [52].

The work of Brüssow and Brüssow reported that medical reports from Britain and Germany on patients suffering from the Russian flu share several characteristics with COVID-19 [4]. Most notable are multisystem affections comprising respiratory, gastrointestinal and neurological symptoms, including loss of taste and smell perception. In COVID19 and unlike in influenza, mortality was seen in elderly subjects, while children were only weakly affected [4].

The Russian flu pandemic claimed the lives of an estimated 1 million humans from a world population of 1.5 billion people and represented thus one of the great epidemics of the 19th century [53]. The pandemic spread was extremely rapid, with a starting point at St Petersburg in December 1889 [53]. The UK and Scottish cities were hit only six weeks later. The mean basic reproduction rate was 2.15, and the highest reproduction rates were observed at Stuttgart, St Petersburg, and Amsterdam [53].

The Russian flu was described as influenza because viruses were still unknown at the time. Since the oldest influenza viruses were isolated and kept as laboratory stocks only since the 1930s, direct evidence for linking influenza viruses with the Russian flu is lacking [4]. In contrast, direct virological proof for the attribution of the Spanish flu from 1918 to 1919 to an influenza virus has been achieved by finding pathological samples and corpses of pandemic victims buried in permafrost soils, followed by reviving this pandemic influenza virus in the laboratory [4].

To address the question of whether the clinical symptoms reported for the Russian flu patients better fit “an influenza virus infection or a trans-species infection h a bovine coronavirus or another infectious agent,” Brüssow and Brüssow used two comprehensive contemporary reports on the Russian flu pandemic from Britain and Germany [4]. According to the British Parsons Report (1891) [54], Brüssow and Brüssow concluded that many observations described in the Parsons report resemble more characteristics of COVID-19 than those of influenza [4]. Notable are light affection in adolescents and age as a risk factor for mortality: “Influenza was a disease especially fatal to elderly persons” [54]. “Pulmonary inflammation was the most frequent cause of death and affected the very old and the previously diseased” [54].

Kousoulis and Tsoucalas [55] also concluded that some characteristics of the 1889 pandemic resemble more coronavirus affection than classical influenza. Further insight is provided by an Encyclopaedia Britannica entry on “Influenza” published in 1911 [56]. According to Encyclopaedia Britannica from 1911, “influenza melancholia is twice as frequent as all other forms of insanity put together. Other common after-effects are weakness or “loss of the special senses, particularly taste and smell” [56]. The German “Verein für Innere Medizin” Report issued in 1892 at Berlin [57] also lists loss of smell and taste.

We have listed above some of the sources we discovered using GNV to confirm the thesis that the Russian flu was the coronavirus infection, i.e. that COVID-19 is not a unique phenomenon. In the following, we begin to show how to use NGram concerning pandemics throughout history and lessons for today. The first example relates to the above symptoms that GNV correctly records, which is the first evidence of the reliability of this approach.

Figure 2 shows the increase in the mention of symptoms “pneumonia; weakness; loss of appetite; headache; bronchitis; febrile temperature; depression and muscular pain” in English book corpora at the time of the outbreak of the Russian flu (1889-1891). We chose the years 1880, 1890, 1900 and 1910 to show the frequencies of mentioning symptoms in the period before the outbreak of the Russian flu and in the period after. Figure 1 indicates that NGram is a reliable tool for monitoring social trends in the past. The section Results and Methods describes the detailed analysis and explanation of the methodology.

**Figure 1.**
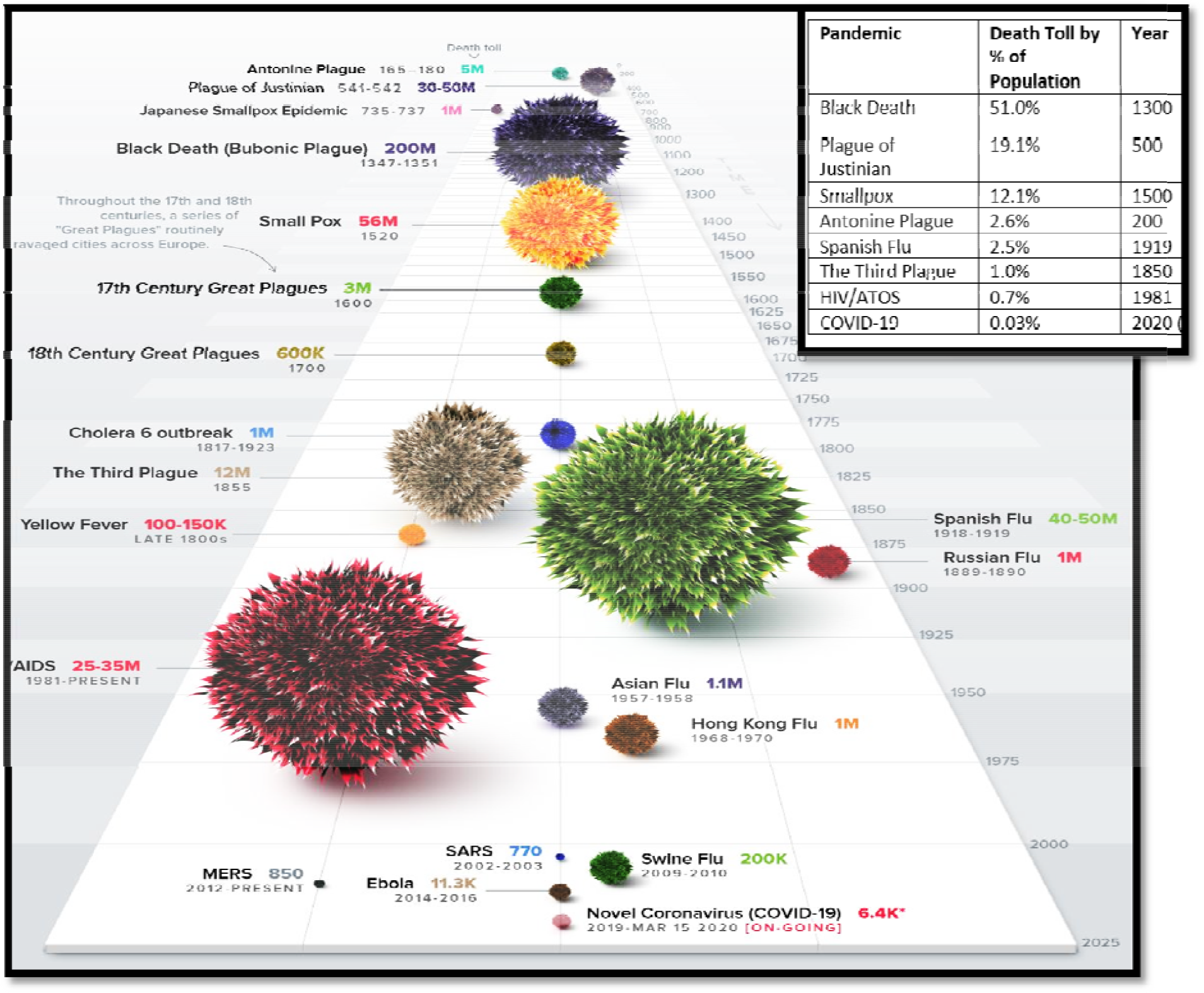
Timeline of historical pandemics. Source: Visual capitalist, CDC, WHO, BBC, Encyclopedia Britannica (https://lider.media/poslovna-scena/svijet/infografika-sve-pandemije-kroz-povijest-130435), edited by author

**Figure 2.**
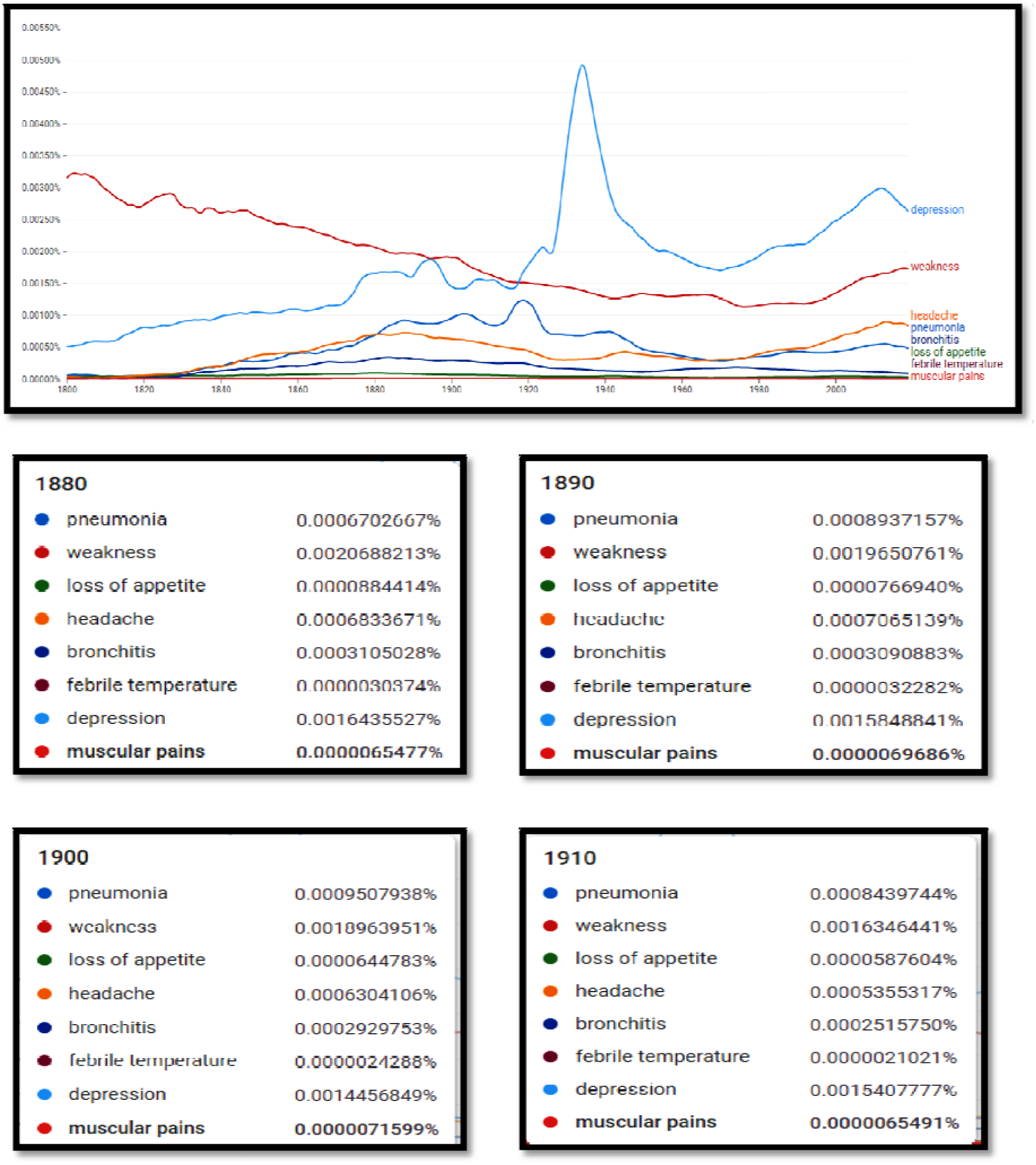
Frequencies for the symptoms “pneumonia; weakness; loss of appetite; headache; bronchitis; febrile temperature; depression and muscular pain” mentioned in newspapers, magazines and books from 1800 to 2019 in the English corpus (Google Ngram) Source: author’s creation based on Google Ngram (http://books.google.com/ngrams)

### Comparison of similarities between COVID-19 and the Russian flu

According to Van Ranst, “incidences like COVID-19 happened all the time, but we did not notice them” - medicine is detecting viruses nowadays more frequently [1]. “If some of these outbreaks, like SARS in 2003, happened one hundred years ago, then it would not have been noticed, and it would be a local outbreak” [1]. In the context of the current pandemic, it is surprising that the COVID-19 virus was sequenced so quickly, especially when considering that one of the most common cold viruses, OC43, had not even been sequenced until 2003 by Mark Van Ranst et al. [1].

It is, of course, difficult to formulate a hypothesis for a microbiological aetiology of a pandemic that occurred 133 years ago, at an epoch when viruses were still unknown. But differentiating an influenza virus infection from a COVID-19 patient purely on the clinical ground is a problematic task for a physician today [4] because the symptoms overlap. As we have already stated, the most important observations of the loss of smell and taste (anosmia and ageusia) were made during the Russian flu pandemic and with COVID-19. Since anosmia and ageusia are now used as relatively reliable clinical diagnostic markers for COVID-19 ^58^, one is tempted to attribute this specific symptom seen in the Russian flu pandemic patients more to a coronavirus than to an influenza virus infection.

According to a thesis from Van Ranst [1] and a reformulated thesis of Telenti et al. [59], the world faced in 1890 a coronavirus pandemic. Due to the mentioned limitations, it is impossible to have medical evidence. Therefore, we have looked for evidence in history using the method of Digital Humanities and GNV below.

Figure 3 shows the rapid increase in the mention of the term “anosmia” (loss of smell) and “ageusia” (loss of taste) in English book corpora at the time of the outbreak of the Russian flu and immediately after it (1889-1891).

**Figure 3.**
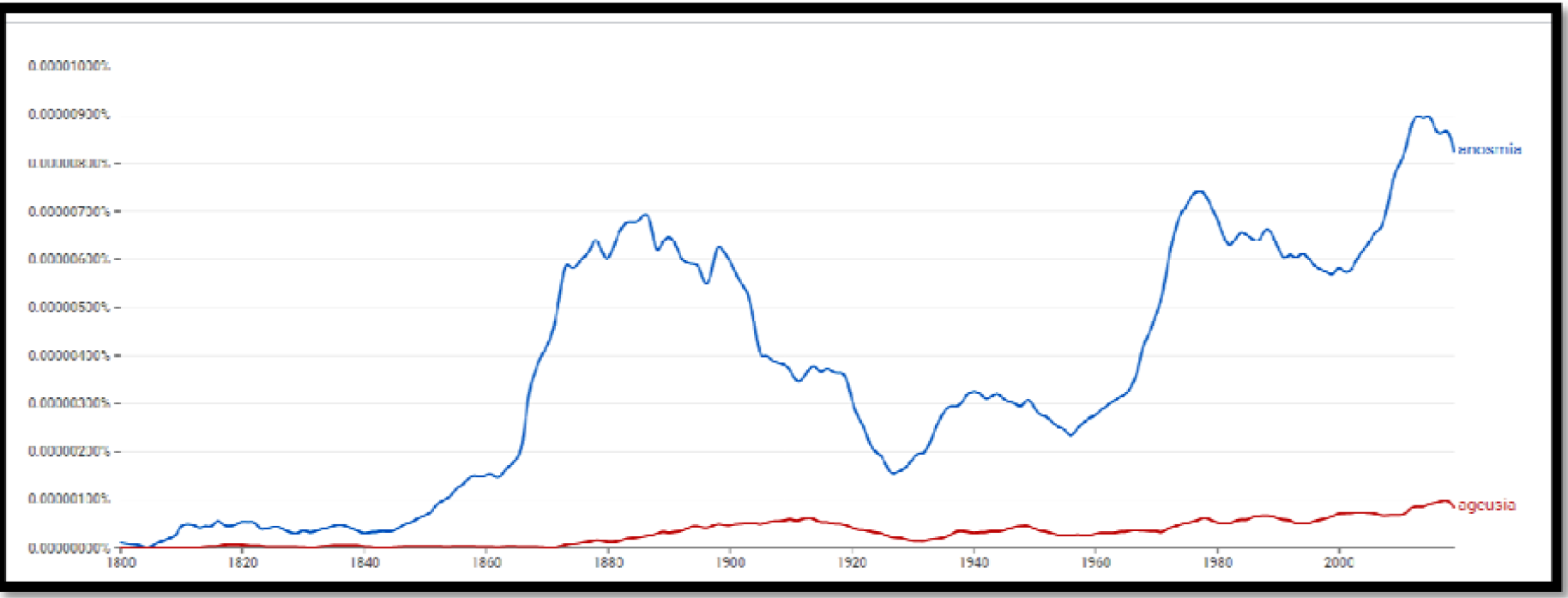
Frequencies for the words “anosmia” and “ageusia” from 1800 to 2019 in the English corpus.

Figure 4 shows the increase in the mention of the term “loss of smell” and “loss of taste” in English book corpora (including newspapers and magazines) at the time of the outbreak of the Russian flu and immediately after it (1889-1891). We can see the same development in German and Russian book corpora (Figure 5 and 6).

**Figure 4.**
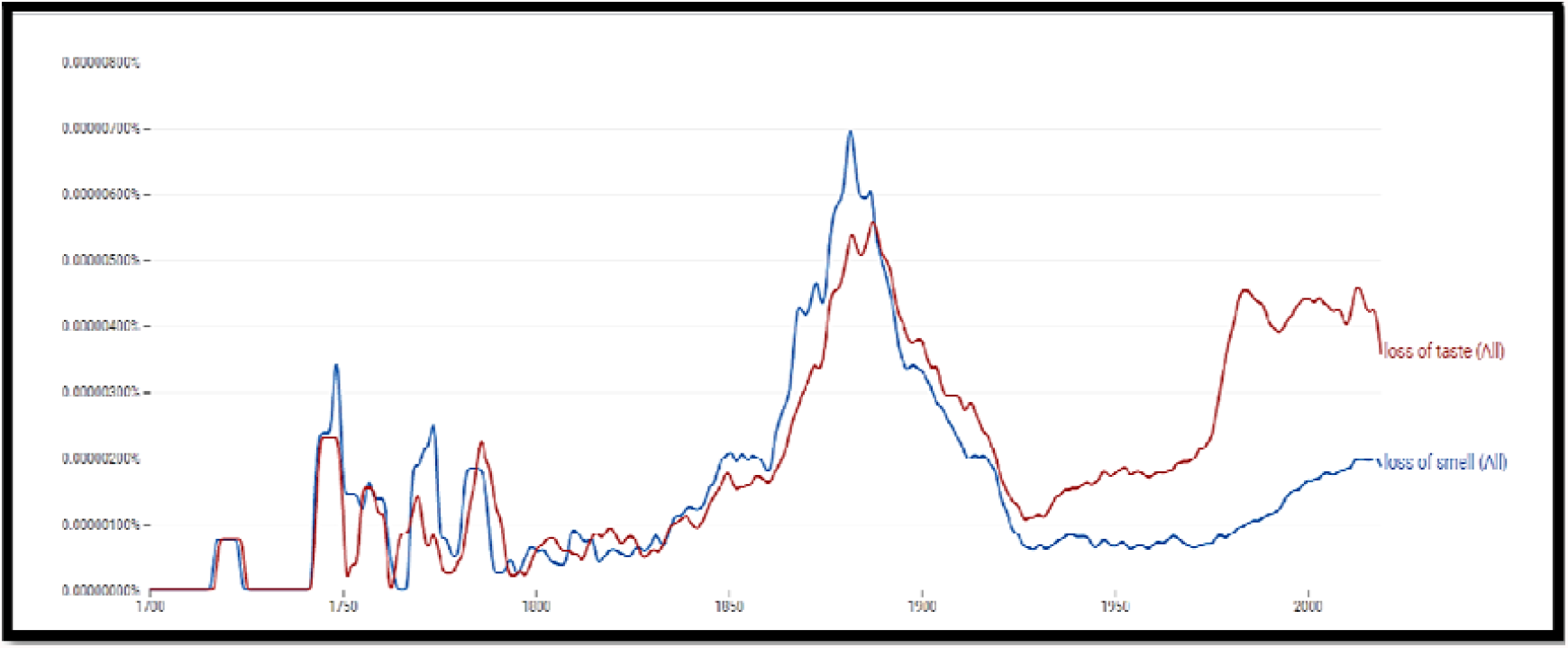
Frequencies for the words “loss of smell” and “loss of taste” from 1700 to 2019 in the English corpus.

**Figure 5.**
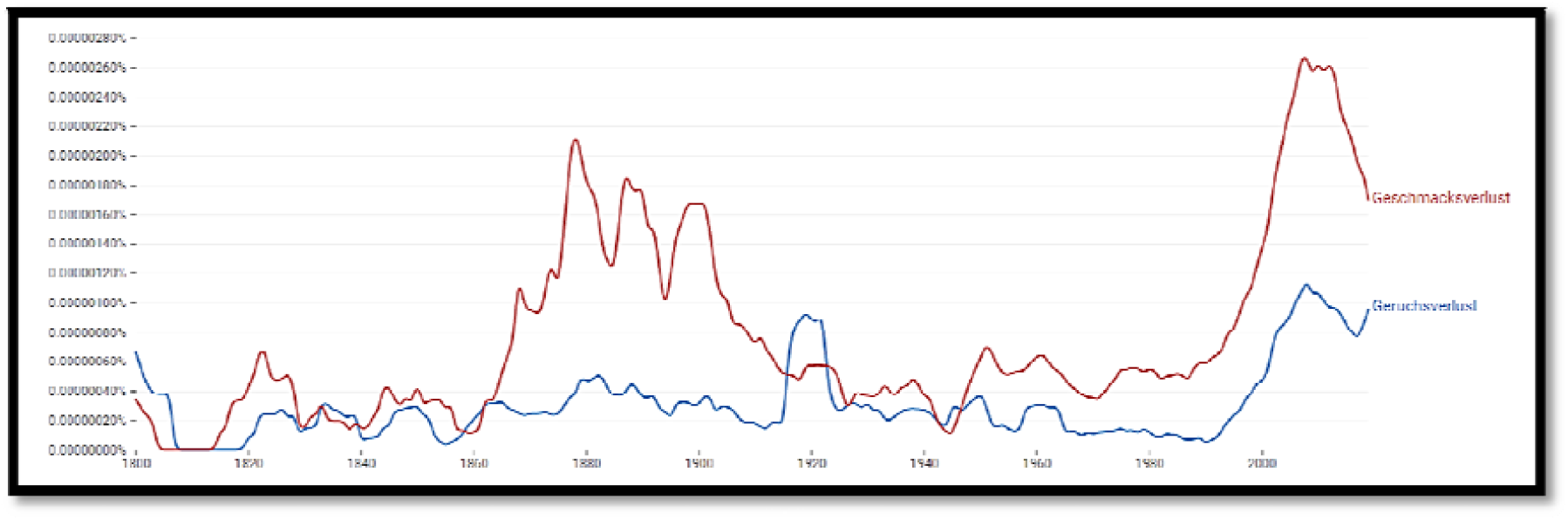
Frequencies for the words “Geruchsverlust” (loss of smell) and “Geschmackverlust” (loss of taste) from 1700 to 2019 in the German corpus.

**Figure 6.**
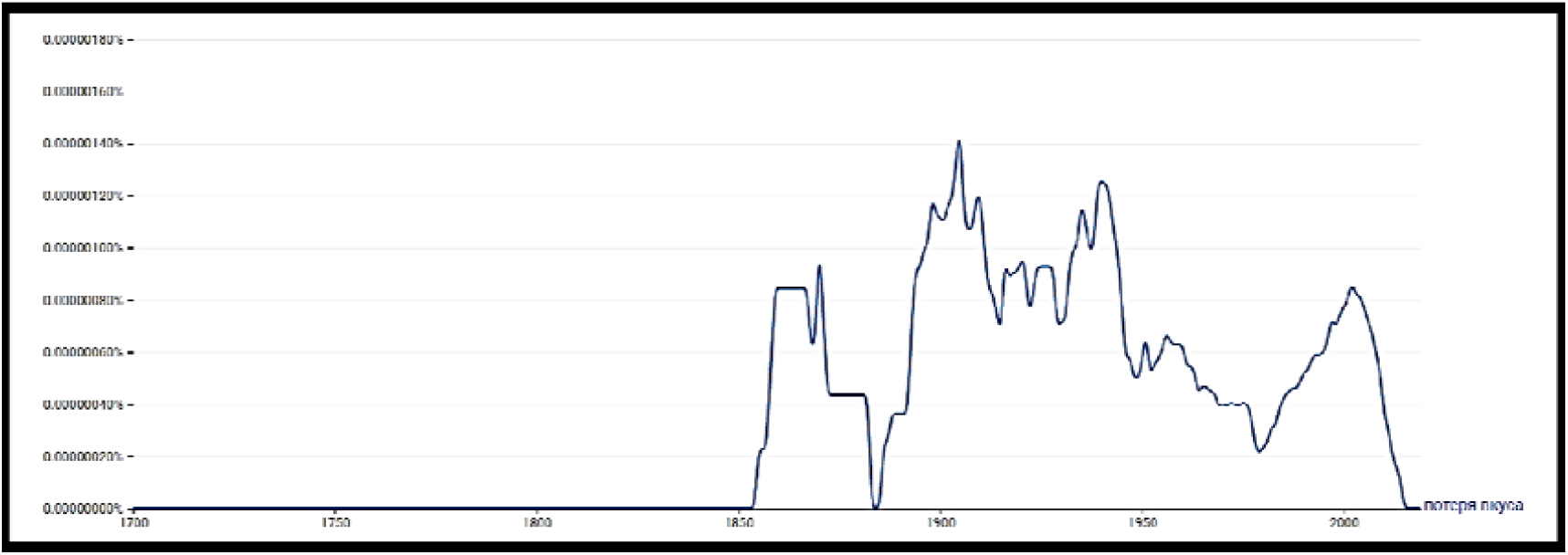
Frequencies for the words “потеря вкуса (loss of taste) “from 1700 to 2019 in the Russian corpus.

In contrast to the German and English one, the Russian corpus (Figure 6) indicates censorship because the terms quickly disappear from the public space after their sudden appearance, i.e. it is no longer mentioned in newspapers or books. The possibility of censorship is also mentioned in the work of Harald Brüssow [3].

The English One Million option allows searches that limit books to 6000 in any given year. Google has made attempts to select books randomly, but at the same time to maintain the subject distributions for each year [21]. Figure 7 also shows, in this case, an apparent increase in the use of the term “loss of smell” in books during the Russian flu.

**Figure 7.**
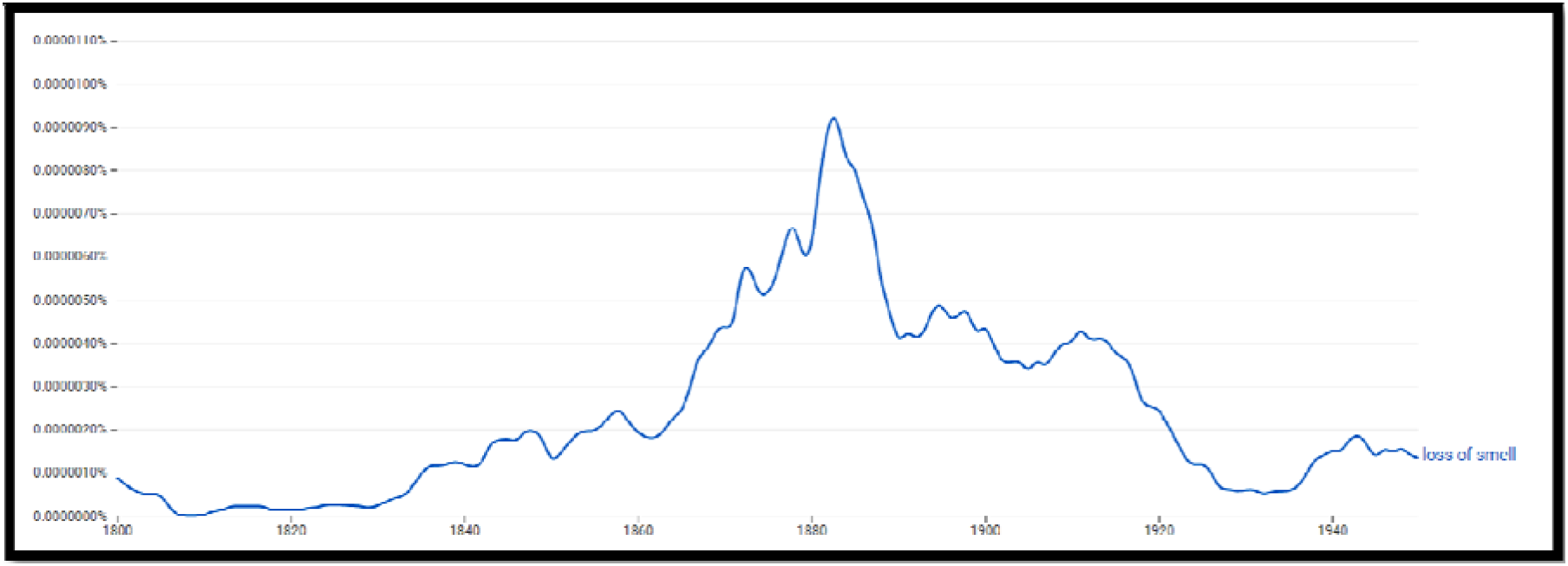
Frequencies for the term “loss of smell” from 1800 to 2019 in the corpus *English One Million*.

Further similarities between Russian flu and COVID-19 are that COVID-19 has, as mentioned, its main fatality in the elderly; this was also noted for the Russian flu pandemic [4]. While the peak mortality in the Russian flu pandemic was with the elderly, substantial mortality was also seen in adults with comorbidity, but children suffered only mild symptoms similar to the current COVID-19 pandemic [60].

In our study, by applying NGram, we also evaluated historical reports from newspapers and scientific and medical journals. GNV recorded more than 600 news articles about the Russian flu from 42 newspapers (Paris - Le Temps, Le Matin, Berlin - Vossische Zeitung, London - The Times, and many Austrian newspapers and medical journals such as The Lancet).

The high attack rate of Russian flu can be read from the newspapers that reported the closure of schools, universities and factories because a large part of the staff fell ill. Reports quoted by the newspapers noted that mortality rates had increased by 30% compared to the same period of the pre-pandemic year.

The past pandemic has elements relevant to the COVID-19 pandemic, showing the measures that we undertake today and the same as they did in 1918 – social distancing, wearing masks, quarantining, and travel restrictions [1]. But just as individuals forget about the past, so do societies [2]. Studying past pandemics shows that the pandemic stops on its own. According to mentioned historical records, a pandemic’s ‘natural’ length is two to five years [61]. In the absence of treatments and a vaccine, both the Russian and the Spanish flu ran and stopped after two to three years. The wearing of masks was during the Spanish flu understood to be of significant importance in preventing infection [62]. However, “herd immunity” was not necessary to stop the pandemic [63].

Despite the similarities, several differences distinguish the COVID-19 situation from the Russian flu. In contrast to its widespread use during the Spanish flu pandemic of 1918, face masks were not used during the Russian flu pandemic [61].

NGram (Figure 8) shows us evidence that during the Russian flu wearing masks was less used than in the period of Spanish flu. This is another proof that NGram correctly records social trends.

**Figure 8.**
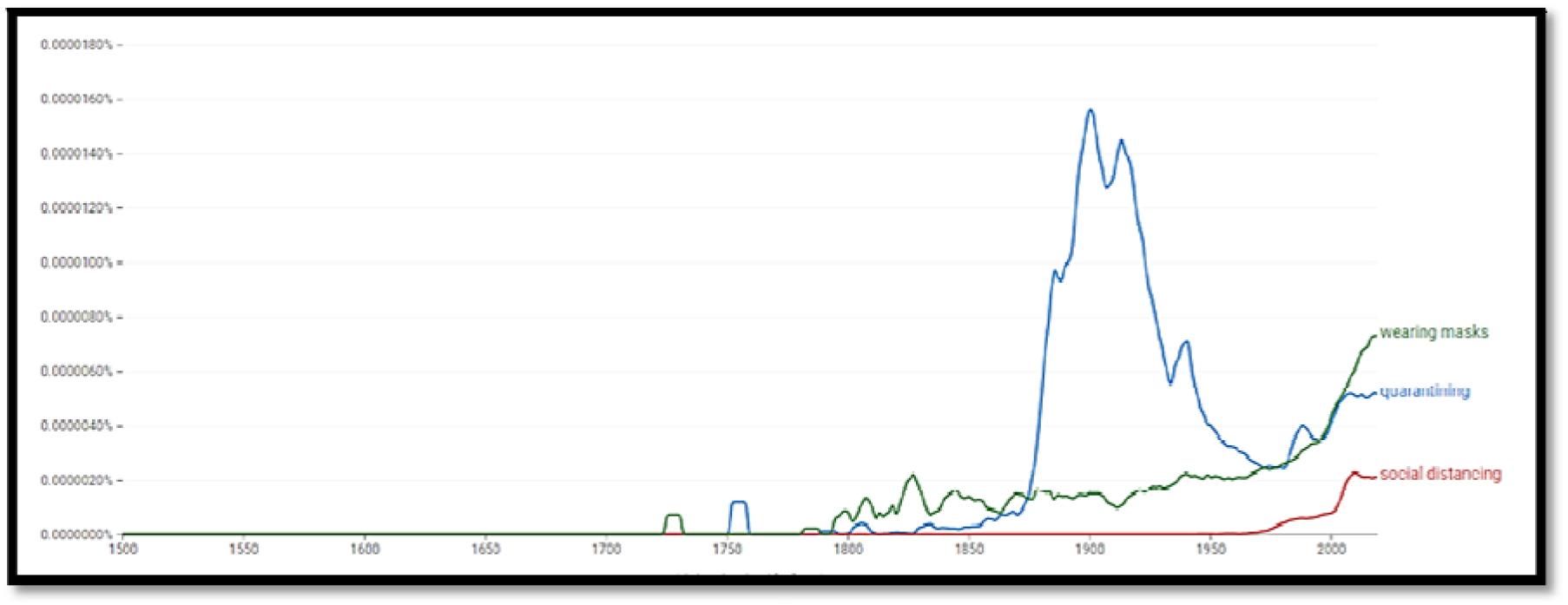
Frequencies for the terms “quarantining “, “social distancing “, and “wearing masks“from 1500 to 2019 in the English corpus.

**Figure 9.**
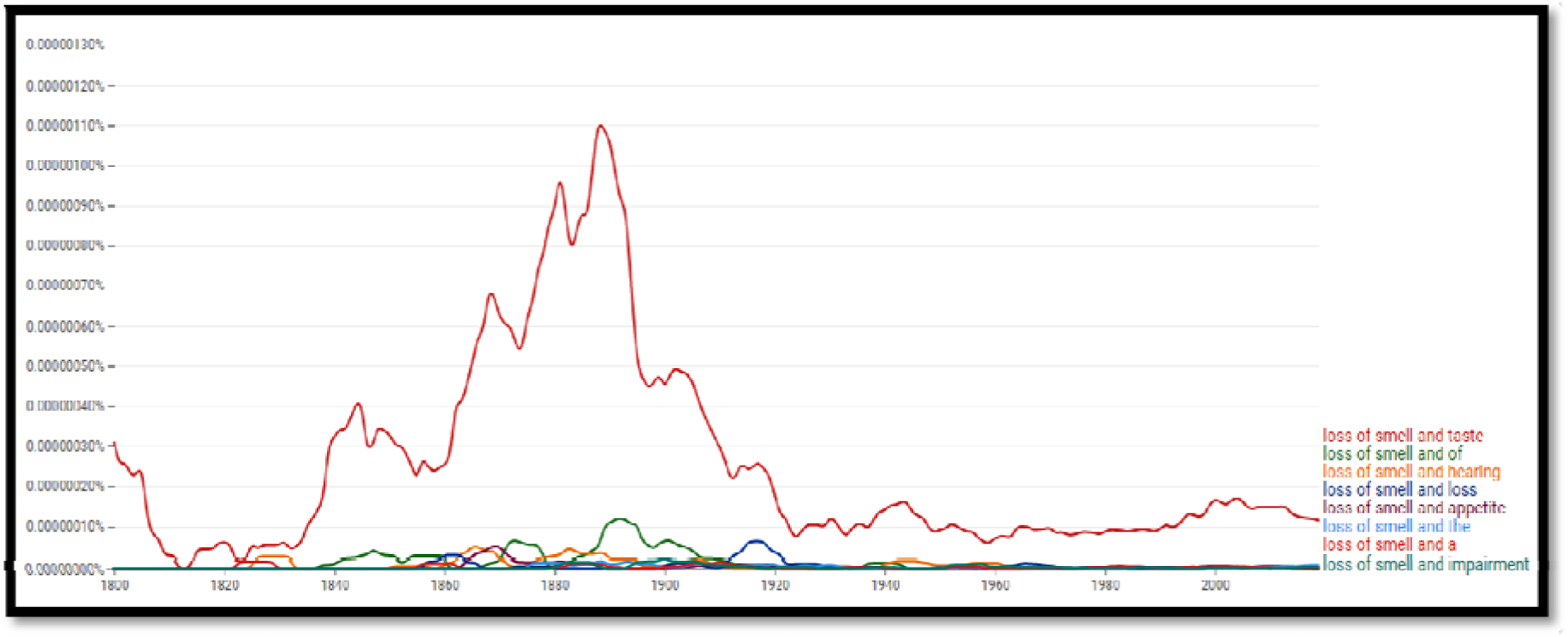
Frequencies for the words “loss of smell and *” from 1800 to 2019 in the English corpus showing most often mentioned followed words using operator “*”.

**Figure 10a:**
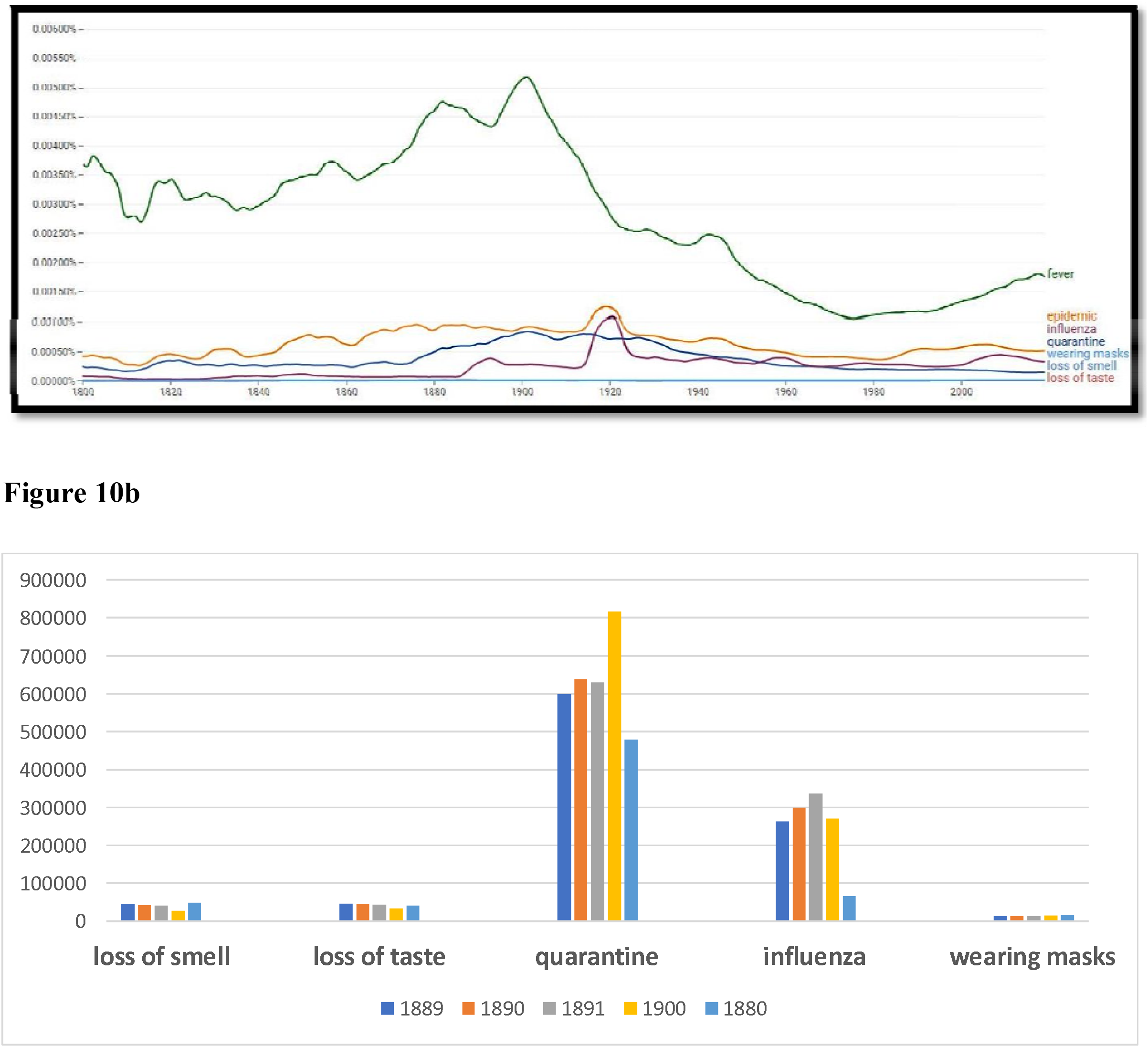
GNV display - Frequencies for the words “fever”, “epidemic”, “influenza”, “quarantine”, “wearing masks”, “loss of smell”, “loss of taste” from 1800 to 2019 in the English corpus.

**Figure 11.**
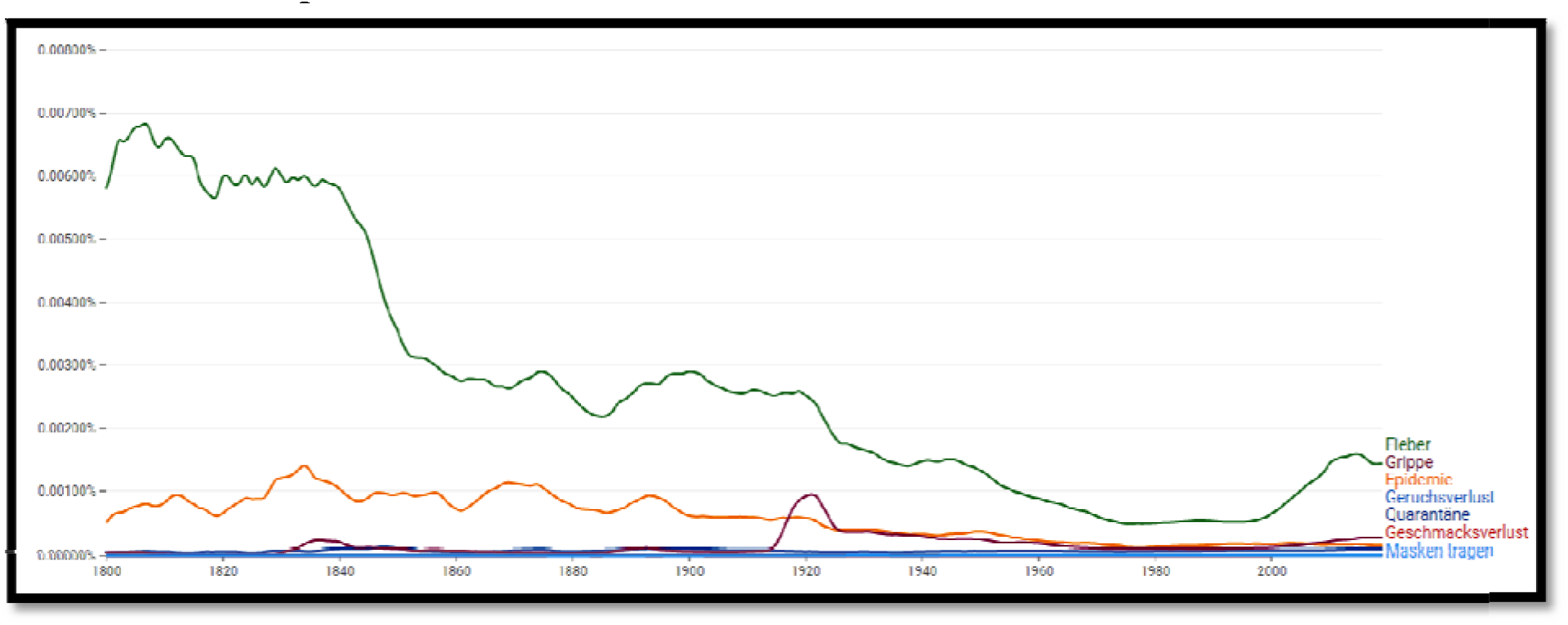
Frequencies for the words “Fieber (fever)”, “Epidemie (epidemic)”, “Grippe (influenza)”, “Quarantäne (quarantine)”, “Masken tragen” (wearing masks), “Geruchsverlust (loss of smell)”, “Geschmacksverlust (loss of taste)” from 1800 to 2019 in the German corpus.

**Figure 12.**
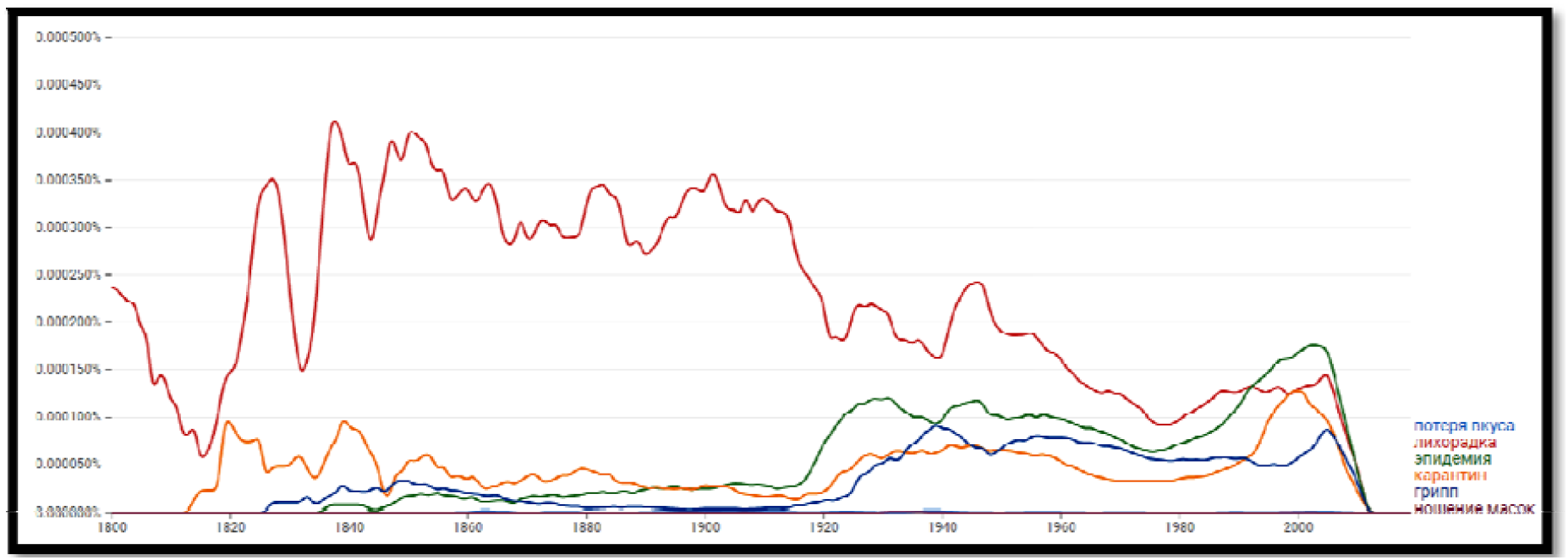
Frequencies for the words “лихорадка” (fiver), “эпидемия” (epidemic), “influenza” (грипп,), “карантин” (quarantine), “ношение масок” (wearing masks), “Потеря обоняния” (loss of smell), “потеря вкуса” (loss of taste) from 1800 to 2012 in the Russian corpus.

The term “social distancing” is a newer word coin, so it is not surprising that it was not mentioned in the 19 century, while in the case of the term “quarantining” we see that the term was intensive mentioned in the middle of the XVIII century (bubonic plague between 1738 and 1740), and that it is intensively mentioned during both the Russian and the Spanish flu. Quarantine was first introduced by Croatia, i.e. Dubrovnik, in the middle of the 14th century [64], but since the printing press was invented in the middle of the 15th century such a record cannot be registered by the NGram (this should be borne in mind in the case of many other discoveries and historical events).

Public health measures during the 1889 pandemic consisted mainly of school closures and hygiene advice (handwashing) that GNV also records [3]. Intensive care medicine was in 1889 practically non-existent, and the best medical advice of the time was early bedrest and antipyretics [3].

Judging by the collective memory of humanity and the insights we have gained using GNV, the virus will undoubtedly weaken over time. The results of GNV show that the pandemic in this decade will turn into an endemic or common cold and will stay with us like other types of flu.

## Methods and results

## Methods

In our work we have used the new updated English corpora (2019) to exploit the advantages of improved OCR and better underlying library and publisher metadata. We chose to work on an English (both British and American) corpus, as it is the most extensive database available so far. For comparison and verification of results, we have also used both the German (2019) and Russian corpora (2012).

The objective was to calculate the ratio of increasing to decreasing trends in the changes in frequencies of the words representing symptoms of the Russian flu to compare similarities between the development of the Russian flu and COVID-19. If the desired term or set of words is entered in the search engine, for example, the word “epidemic”, the graphic display on the Y-axis shows what percentage of words in the selected corpus over the years (X-axis) make up the word.

It is important to emphasise that the smoothing factor “3” we use in the paper shows the average for each year, considering the three previous and three upcoming years. The validity of the data obtained is guaranteed by normalising the data with the number of published books each year [14].

As previously mentioned, GNV provides five operators that the researcher can use to combine n-grams: “+, -, /, *, :” With the “wild card”, a searcher can ask for information that is not predefined by other search keywords. That can lead to an exploration of hidden patterns [10]. The wild card can be applied to the next adjacent word and different patterns. When the researcher puts a “*” in place of a word, the Ngram Viewer will display the top ten substitutions. For instance, to find the most popular words following “University of” search for “University of *” [16].

For our study, this operator is helpful in the context that it shows that the term “loss of smell” is most often mentioned in combination with the term “loss of taste”. In addition, we see that both terms are used frequently during the Russian flu.

Below the graph, GNV shows year ranges for query terms, and by clicking on those the query is directly submitted to Google Books. It is important to note here that one can choose between newspapers, magazines and books.

### 3.2 Results

The frequency of the words “loss of smell” and “loss of taste” rapidly increased in the English corpus during the Russian flu from 1899 to 1891. In the case of symptom “*loss of taste,”* the frequency rose from 0.0000040433 % in 1880 to 0.0000047123 % in 1889. One can also notice that the mention of this symptom fell sharply after the pandemic stopped in 1900 (0.0000033861%). In the case of symptom “*loss of smell,”* the frequency decreased from 0.0000043904 % in 1889 to 0.0000028211 % in 1900.

In the German corpus the frequency for “Geschmacksverlust” (loss of taste) rose from 0.0000014463 % in 1880 to 0.0000018015 % in 1889 and decreased rapidly after the pandemic (1900 = 0.0000016600 %). The most rapid change in the German corpus between the years 1890 and 1900 can be noted at the term “Epidemie” (epidemic) (1890 = 0.0008011005 %; 1900 = 0.0005970067 %).

In the Russian corpus the frequency for “loss of taste” rose from 0 % in 1880 to 0.0000004682 % in 1889 and decreased rapidly after the pandemic (1900 = 0.0000011834 %). The frequency for “loss of smell” rose from 0. 000000000 % in 1880 to 0.0000005041 % in 1889 and decreased rapidly after the pandemic (1900 = 0.0000001579 %).

The comparison presented in Table 4. clearly shows that all the three corpora (English, German and Russian corpus) we used for the analysis show that the symptoms of “loss of taste” before and after the outbreak of the Russian flu pandemic were mentioned in the literature, newspapers and magazines to a much lesser extent then it was during the pandemic. The same is noticeable in almost all other symptoms and social trends.

**Table 1.** Frequencies for the words “fever”, “epidemic”, “influenza”, “quarantine”, “wearing masks”, “loss of smell”, “loss of taste” from 1800 to 2019 in the English corpus (in %)

|  |  |  |  |  |  |
| --- | --- | --- | --- | --- | --- |
| Russian flu | 1880 | 1889 | 1890 | 1891 | 1900 |
| loss of smell | 0.0000049357 | 0.0000043904 | 0.0000042023 | 0.0000040161 | 0.0000028211 |
| loss of taste | 0.0000040433 | 0.0000047123 | 0.0000044648 | 0.0000043490 | 0.0000033861 |
| fever | 0.0045785303 | 0.0044563019 | 0.0044123274 | 0.0043885018 | 0.0051403633 |
| epidemic | 0.0008526997 | 0.0008944025 | 0.0008965982 | 0.0009177670 | 0.0008983375 |
| quarantine | 0.0004794893 | 0.0005985671 | 0.0006383930 | 0.0006295467 | 0.0008166632 |
| influenza | 0.0000663529 | 0.0002639855 | 0.0002983151 | 0.0003359815 | 0.0002702021 |
| wearing masks | 0.0000015962 | 0.0000013616 | 0.0000013950 | 0.0000013922 | 0.0000015227 |

**Table 2.** Frequencies for the words “Fieber (fever)”, “Epidemie (epidemic)”, “Grippe (influenza)”, “Quarantäne (quarantine)”, “Masken tragen” (wearing masks), “Geruchsverlust (loss of smell)”, “Geschmacksverlust (loss of taste)” from 1800 to 2019 in the German corpus (in %)

| Russische Grippe | 1889 | 1890 | 1891 | 1900 | 1880 |
| --- | --- | --- | --- | --- | --- |
| Geruchsverlust | 0.0000004145 | 0.0000003559 | 0.0000003501 | 0.0000003071 | 0.0000003501 |
| Geschmacksverlust | 0.0000017517 | 0.0000018015 | 0.0000014463 | 0.0000016600 | 0.0000014463 |
| Fieber | 0.0024394190 | 0.0025105012 | 0.0026102443 | 0.0029010263 | 0.0026102443 |
| **Epidemie** | 0.0007245716 | 0.0008011005 | 0.0008347561 | 0.0005970067 | 0.0008347561 |
| **Quarantäne** | 0.0000705233 | 0.0000763311 | 0.0000766396 | 0.0000863325 | 0.0000766396 |
| **Grippe** | 0.0000711845 | 0.0000807742 | 0.0000877451 | 0.0000481621 | 0.0000877451 |
| **Masken tragen** | 0.0000006314 | 0.0000004534 | 0.0000003115 | 0.0000012416 | 0.0000003115 |

**Table 3.** Frequencies for the words “лихорадка” (fiver), “эпидемия” (epidemic), “influenza” (грипп,), “карантин” (quarantine), “ношение масок” (wearing masks), “Потеря обоняния” (loss of smell), “потеря вкуса” (loss of taste) from 1800 to 2012 in the Russian corpus (in %)

|  |  |  |  |  |  |
| --- | --- | --- | --- | --- | --- |
| **Russian flu** | **1889** | **1890** | **1891** | **1900** | **1880** |
| **Потеря обоняния** | 0.0000005041 | 0.0000005041 | 0.0000005041 | 0.0000001579 | 0.000000000 |
| **потеря вкуса** | 0.0000004682 | 0.0000004682 | 0.0000006787 | 0.0000011834 % | 0.000000000 |
| **лихорадка** | 0.003102872 | 0.003102872 | 0.0002993711 | 0.0003607911 | 0.0003471586 |
| **эпидемия** | 0.0000247684 | 0.0000247684 | 0.0000278790 | 0.0000251270 | 0.0000191910 |
| карантин | 0.0000277171 | 0.0000277171 | 0.0000254140 | 0.0000297613 | 0.0000473150 |
| грипп | 0.0000065101 | 0.0000065101 | 0.0000057692 | 0.0000044993 | 0.000052766 |
| ношение масок | 0.0000000000 | 0.0000000000 | 0.0000000000 | 0.0000000000 | 0.0000000000 |

**Table 4.** Comparison for the symptom “loss of taste” in the English, German and Russian book corpus (in %)

| loss of taste | 1880<br>↓ | 1889<br>↑ | 1890<br>↑ | 1891<br>↑ | 1900<br>↓ |
| --- | --- | --- | --- | --- | --- |
| English corpus | 0.0000040433 % | 0.0000047123 | 0.0000044648 | 0.0000043490 | 0.0000033861 |
| German corpus | 0.0000014463 | 0.0000017517 | 0.0000018015 | 0.0000014463 | 0.0000016600 |
| Russian corpus | 0.0000000000 | 0.0000004682 | 0.0000004682 | 0.0000006787 | 0.0000011834 |

The frequency of the words “fever”, “epidemic”, “influenza”, “quarantine”, “wearing masks”, “loss of smell”, “loss of taste” increased rapidly during the Russian flu from 1899-1891, which is especially noticeable in the German and Russian book corpus. In the case of symptom “*loss of taste” in the English corpus*, the frequency rose from 0.0000040433 % in 1880 to 0.0000047123 % in 1889. One cannot but notice that the mention of this symptom fell sharply after the pandemic stopped in 1900 (0.0000033861%).

In the Russian corpus, the frequency rose from 0. 000000000 % in 1880 to 0.0000004682 % in 1889 and decreased rapidly after the pandemic (1900 = 0.0000011834 %). In the German corpus the frequency rose from 0.0000014463 % in 1880 to 0.0000018015 % in 1889 and decreased rapidly after the pandemic (1900 = 0.0000016600 %). This proves our thesis that GNV is a reliable tool for monitoring social trends during pandemics and a very useful window into history.

Of the other social trends we have analysed using GNV, we would highlight the terms: “economic crisis”, “unemployment”, and “hunger”. None of these terms shows a significant deviation in frequencies compared to the period immediately before and after the epidemic. We conclude that looking from the historical point of view (GNV as the window of history) that a significant crisis does not need to occur after the COVID-19 pandemic.

## Limitations

The possibilities and limitations of using the GNV for research have been controversially discussed [20]. Although many Google Ngram studies indicate scientific recognition, several papers address methodological issues [65]. The data set from GNV has been criticised for its reliance on inaccurate OCR, an overabundance of scientific literature, and for including large numbers of incorrectly dated and categorised texts [66]. Because of these errors, and because it is uncontrolled for bias, according to Zhang, it is risky to use this corpus to study language or test theories [67]. Since the data set does not include metadata, it may not reflect the general linguistic or cultural change and can only hint at its effect [20].

The main points of criticism relate to insufficient OCR, particularly concerning semantic scanning errors (which can affect words such as fail and sail due to similarities in the letters “f” and “s”) [66], and messy metadata that may lead to the display of word frequencies in the wrong or unrelated time intervals [65]. The last criticism is that the percentage considers published manuscripts regardless of their importance [22].

Hilpert and Gries [68] warn that a statistical measure that would help determine if the observed frequencies differ from the mean more than expected should be incorporated in more complex studies. Mayer-Schönberger and Cukier express concern about machines replacing human activities and decision-making [20]. Boyd and Crawford also raise critical questions about big data: “Will large-scale search data help us create better tools, services, and public goods? Or will it usher in a new wave of privacy incursions and invasive marketing? (…) The era of Big Data has only just begun, but it is already important that we start questioning the assumptions, values, and biases of this new wave of research “[69].

Several authors have problematised the GNV corpus and raised doubts about it representing natural language and its development over time [66]. Chumtong and Kaldewey highlight that what makes the GNV a valuable research tool is not primarily its accuracy but rather its potential for “quick-and-dirty heuristic analysis” [18]. Davis [70] recognised the dataset as remarkable, but perceived the interface as too simplistic. He claimed it did not allow for collocations in searches, searching by wildcards and meaningful use of parts of speech.

It also appears that GNV does not consider the different contexts in which the analysed words are set in, and contexts carry the meaning the cause of which we are unable to determine. The fact that the frequency of a word rises does not necessarily mean that the concept is valued more, but that it is discussed extensively [13]. The GNV enables viewing the excerpts from which the analysed words come; however, as collecting such data has not been automated yet, and would have to be done manually for all words in millions of contexts, it seems implausible to incorporate such information into the study, even if for reasons of time and space [13]. Needless to say, either an individual or a larger team cannot study any of the corps manually.

According to Zieba, the usage of GNV should be limited to uncomplicated studies related to word frequency. It cannot be treated as the only tool in researching complex socio-cultural transformations [13]. However, with careful analysis of the results, the GNV does potentially improve our understanding of cultural and linguistic trends over time. With Google making its datasets available, more complex text mining tools can study the ever-growing corpus [21]. Compared to the 2009 versions, the 2012 and 2019 versions have more books, improved OCR, improved library and publisher metadata [16]. According to Zieba, even if we consider the imperfections of OCR, GNV still seems to put socio-cultural research in a context whose significance is hard to question, especially if carried out cautiously and conscientiously [13].

Lakoff agrees that even though the presence of most words and the changes in their frequency does not tell much about the values ascribed to certain phenomena, it may be a sign of recognition of a problem [71]. Younes and Reips propose how to address these concerns by introducing several methodological procedures such as cross-validations via the examination of different language corpora, the use of word inflexions and synonyms, as well as the use of a newly-developed standardisation procedure that all aim at increasing the reliability of GNV studies [20].

According to Solovyev et al., there are several ways to make the GBN corpus results more reliable [72]. On the one hand, it is impossible to correct all its errors, and on the other, perfectionism should be avoided in this field since no one knows what an ideal corpus would be like [72]. The first one is to use all possible support data extracted from the corpus and use synonyms [20]. Younes recommends studying each word and its three synonyms selected from the relevant dictionaries of synonyms [20]. Sometimes it is pertinent to perform comparative studies and see how the same or close in meaning terms are used in different corpora presented in GNV [72]. The second way to enhance the results is to pre-process the GNV raw data. Solovyev et al. show that the GNV corpus can be regarded as representative for the following reasons. It is the most extensive corpus ever existed, including texts of various types and genres written by people of different ages, sex and with diverse backgrounds. Such diversity of texts, their length and size serve as a solid empirical foundation for linguistic and related studies [72].

## Conclusion

This paper showed that the Google Ngram (GNV) can give us useful insights into the history of pandemics, and that the tools of Digital Humanities can discover hidden patterns in history. With the help of GNV we have analysed the epidemiological literature on the Russian flu pandemic development for hints on how the COVID-19 might develop in the following years. We showed evidence that the COVID-19 is not a unique phenomenon because the Russian flu was the coronavirus infection. We are aware that it is difficult to formulate a hypothesis for a microbiological aetiology of a pandemic that occurred 133 years ago, but differentiating an influenza virus infection from a COVID-19 patient purely on the clinical ground is even today for a physician a difficult task because the symptoms are overlapping.

The most important observation of similarities between the Russian flu pandemic and COVID-19 is the loss of smell and taste (anosmia and ageusia). The basic methodology concept was to calculate the ratio of increasing to decreasing trends in the changes in frequencies of the selected words representing symptoms of the Russian flu and COVID-19. We chose to work on an English (both British and American) corpus, as it is the largest database available so far. For comparison purposes and verification of results, we also used German corpora (2019) and Russian corpora (2012). To standardise the data, we requested the data from 1800 to 2019, and focused on the ten years before, during and after the outbreak of the Russian flu. We compared this frequency index with “non-epidemic periods” to prove the signification of results and test the model’s analytical potential.

According to our study, the GNV clearly shows the influence that social changes have on word frequency. The frequency of the words “fever”, “epidemic”, “influenza”, “quarantine”, “wearing masks”, “loss of smell”, “loss of taste” increased rapidly during the Russian flu from 1899-1891, which is especially noticeable in the German and Russian book corpus. In case of the symptoms that are undoubtedly proven to be key symptoms of COVID-19 - “loss of taste” and “loss of smell” - during the Russian flu in English corpus, the frequency rose from 0.0000040433 % in 1880 to 0.0000047123 % in 1889. It is also observed that the mention of this symptom fell sharply after the pandemic stopped in 1900 (0.0000033861%). In the Russian corpus, the frequency rose from 0 % in 1880 to 0.0000004682 % in 1889 and decreased rapidly after the pandemic (1900 = 0.0000011834 %). In the German corpus the frequency rose from 0.0000014463 % in 1880 to 0.0000018015 % in 1889 and decreased rapidly after the pandemic (1900 = 0.0000016600 %).

Of the other social trends we have analysed using GNV, we would highlight the terms: “economic crisis”, “unemployment”, and “hunger”. None of these terms shows a significant deviation in frequencies compared to the period immediately before and after the epidemic. We conclude that a historical perspective shows that a significant crisis does not need to occur after the COVID-19 pandemic.

This result proves our thesis that GNV is a reliable tool for monitoring social trends during pandemics and a very useful window into history. Judging by the collective memory of humanity and the insights we have gained using GNV, the virus will certainly weaken over time. The results of GNV show that the pandemic in this decade will turn into an endemic or common cold and will stay with us like other types of flu.

This study has also showed how to overcome the binderies between the social sciences and the humanities. The results of this study open a discussion on the usefulness of the GNV insights possibilities into past socio-cultural development, i.e. epidemics and pandemics that can serve as lessons for today. We have showed hidden patterns of conceptual trends in history and their relationships with current development in the case of the pandemic COVID-19. Despite the numerous indications we have demonstrated, we are aware that the hypothesis still cannot be confirmed, and that it is necessary to require further historical and medical research. The main challenge was to correctly interpret patterns discovered by digital analysis and discern correlations, causes and relations from historical events with current development.

The benefit of this method with an approach to Digital Humanities, particularly GNV, could help complement historical medical records, which are often woefully incomplete. However, this method has serious limitations and can be useful only under cautious handling and testing. The GNV will find application in many research areas in humanities and social sciences in future. Despite its limitations, the GNV research based on an over 500 billion word corpus is prone to produce valuable results when approached with great care and consideration according to the restrictions brought by this method.

## Data Availability

All data produced in the present work are contained in the manuscript

https://books.google.com/ngrams

## References

1 Why history suggests Covid-19 is here to stay, https://ec.europa.eu/research-and-innovation/en/horizon-magazine/qa-why-history-suggests-covid-19-here-stay [accessed 20.12.2021]

2 Halbwachs, M. On Collective Memory. Trans. Lewis A. Coser. Chicago: University of Chicago, 1992.

3 Harald Brüssow. (2021). What we can learn from the dynamics of the 1889 ‘Russian flu’ pandemic for the future trajectory of COVID-19, Microbial Biotechnologie, Volume 14, Issue 6, 2021, Pages 2244–2253, https://doi.org/10.1111/1751-7915.13916

4 Harald Brüssow, Lutz Brüssow, Clinical evidence that the pandemic from 1889 to 1891 commonly called the Russian flu might have been an earlier coronavirus pandemic, Microbial Biotechnologie, Volume 14, Issue 5, 2021, Pages 1860–1870, https://doi.org/10.1111/1751-7915.13889

5 Tado Juric (2022). Google Trends as a Method to Predict New COVID-19 Cases and Socio-Psychological Consequences of the Pandemic, Athens Journal of Mediterranean Studies - Volume 8, Issue 1, 2021, 67–92, https://www.athensjournals.gr/mediterranean/2022-8-1-4-Juric.pdf

6 Rojas Castro A (2017). Big Data in the Digital Humanities. New Conversations in the Global Academic Context, AC/E Digital Culture 2017 Annual Report, 62–71, http://dx.doi.org/10.17613/M6434X

7 Ward and Barker, 2013, “Undefined by Data: A Survey of Big Data Definitions”

8 Preeti Oza: Digital Humanities-An Introduction in: Gurudutta Pradeep Japee, Preeti Oza: Multidimensionality of the Concept & Function of Digital Publisher, Apple Books 2020

9 Burdick A, Drucker J, Lunenfeld P, Presner T and Jeffrey S: Digital_Humanities, The MIT Press 2012, ISBN: 9780262528863

10 Shai Ophir, (2016) Big data for the humanities using Google Ngrams: Discovering hidden patterns of conceptual trends, First Monday, Volume 21, 7–4, 2016, https://firstmonday.org/ojs/index.php/fm/article/download/5567/5535, DOI: https://doi.org/10.5210/fm.v21i7.5567

11 Bruno Latour, Rematerializing Humanities Thanks to Digital Traces, Digital Humanities 2014 - Opening Night Sciences Paris, https://www.youtube.com/watch?v=4L2zRoKS0IA&ab_channel=UNILUniversit%C3%A9deLausanne [accessed 23.12.2021]

12 Antonio Rojas Castro (2017) Big Data in the Digital Humanities. New Conversations in the Global Academic Context, AC/E Digital Culture 2017 Annual Report, 62–71, http://dx.doi.org/10.17613/M6434X

13 Anna Zieba (2018) Google Books Ngram Viewer in socio-cultural research. DOI: 10.2478/rela-2018-0015

14 Michel, J.B., Shen, Y.K., Presser Aiden, A., Veres, A., Gray, M.K., Brockman, W., Pickett, J.P., Hoiberg, D., Clancy, D., Norvig, P., Orwant, J., Pinker, S., Nowak, M.A. i Lieberman A. E. (2011) Quantitative Analysis of Culture Using Millions of Digitized Books. Science, 331 (6014), 176–182, doi:10.1126/science.1199644

15 Lieberson, Stanley and Joel Horwich. “Implication analysis: a pragmatic proposal for linking theory and data in the social sciences.” Sociological Methodology 38 (December 2008): 1–50. 16 Google NGram View, https://books.google.com/ngrams.

16 Lin, Yuri et al.. (2012): “Syntactic Annotations for the Google Books Ngram Corpus”. Proceedings of the 50th Annual Meeting of the Association for Computational Linguistics. Jeju, Republic of Korea, pp. 169–174.

17 Jason Chumtong, David Kaldewey, Beyond the Google Ngram Viewer: bibliographic databases and journal archives as tools for the quantitative analysis of scientific and meta-scientific concepts, FIW Working paper 08, Bonn, 2017, ISBN 978-3-946306-07-8

18 Michel, J.B., Shen, Y.K., Presser Aiden, A., Veres, A., Gray, M.K., Brockman, W., Pickett, J.P., Hoiberg, D., Clancy, D., Norvig, P., Orwant, J., Pinker, S., Nowak, M.A. i Lieberman A. E., Supporting Online Material for Quantitative Analysis of Culture Using Millions of Digitized Books, www.sciencemag.org/cgi/content/full/science.1199644/DC1

19 Younes N, Reips U-D (2019) Guideline for improving the reliability of Google Ngram studies: Evidence from religious terms. PLoS ONE 14(3): e0213554. https://doi.org/10.1371/journal.pone.0213554

20 University of London, An introduction to text mining, https://port.sas.ac.uk/mod/book/view.php?id=554&chapterid=331

21 Gilles Kratzer, (2019) Google Ngram, https://gilleskratzer.netlify.app/post/ngram/ [accessed 23.12.2021]

22 Juric, Tado (2021). Medical brain drain from South-eastern Europe: using digital demography to forecast health worker emigration, Journal of Medical Internet Research JMIRx Med, http://dx.doi.org/10.2196/30831

23 Marziah Karch (2021), How to Use the Ngram Viewer Tool in Google Books, https://www.lifewire.com/google-books-ngram-viewer-1616701 [accessed 23.12.2021]

24 Lucija Kardaš, Uporaba Google Ngrama u društvenim znanostima, Masterthesis, Zagreb: Hrvatsko katolicko sveucilište, 2020. urn:nbn:hr:224:693619

25 http://storage.googleapis.com/books/ngrams/books/datasetsv2.html. [accessed 23.12.2021]

26 Berry D M, (2012: 1), The Social Epistemologies of Software, A Journal of Knowledge, Culture and Policy, Volume 26, 2012, https://doi.org/10.1080/02691728.2012.727191

27 Rutten, B P F, Hammels C, N. Geschwind, C. Menne-Lothmann, E. Pishva, K. Schruers, D. van den Hove, G. Kenis, J. van Os, M. Wichers (2013: 40), Resilience in mental health: linking psychological and neurobiological perspectives, Acta Psychiatrica Scandinavica, https://doi.org/10.1111/acps.12095

28 Michalski B, Krishnamoorthy M, Lau T Y (2012:1), Temporal Analysis of Literary and Programming Prose, https://www.researchgate.net/publication/221663041_Temporal_Analysis_of_Literary_and_Programming_Prose

29 Newberry, M.G., Ahern, C. A., Clark, R. i Plotkin, J.B. (2017) Detecting evolutionary forces in language change. Nature Research, 00(0)

30 Greenfield, P.M. (2013) The Changing Psychology of Culture From 1800 Through 2000. Psychological Science, 20 (10)

31 Acerbi A., Lampos V., Garnett P. i Bentley R.A. (2013) The Expression of Emotions in 20th Century Books. PLOS ONE, 8(3)

32 Twenge J. M., Campbell W. K., & Gentile B. (2012b). Male and female pronoun use in US books reflects women’s status, 1900–2008. Sex Roles, 67(9–10), 488–493

33 Mohammad S. M. (2012). From once upon a time to happily ever after: Tracking emotions in mail and books. Decision Support Systems, 53(4), 730–741.

34 Roivainen E. (2015). Personality adjectives in twitter tweets and in the Google books corpus. An analysis of the facet structure of the openness factor of personality. Current Psychology, 34(4), 621–625.

35 Virues-Ortega J., & Pear J. J. (2015). A history of “behavior” and “mind”: Use of behavioral and cognitive terms in the 20th century. The Psychological Record, 65(1), 23–30.

36 Rossi E., Mortimer J., & Rossi K. (2013). Therapeutic hypnosis, psychotherapy, and the digital humanities: The narratives and culturomics of hypnosis, 1800–2008. American Journal of Clinical Hypnosis, 55(4), 343–359. pmid:23724569

37 Mooijman M., Meindl P., Oyserman D., Monterosso J., Dehghani M., Doris J. M., et al. (2018). Resisting temptation for the good of the group: Binding moral values and the moralization of self-control. Journal of Personality and Social Psychology, 115(3), 585–599. pmid:28604018

38 Roivainen E. (2014). Changes in word usage frequency may hamper intergenerational comparisons of vocabulary skills: An Ngram analysis of wordsum, WAIS, and WISC test items. Journal of Psychoeducational Assessment, 32(1), 83–87

39 Kesebir S., & Kesebir P. (2017). A growing disconnection from nature is evident in cultural products. Perspectives on Psychological Science, 12(2), 258–269. pmid:28346112

40 Grossmann I., & Varnum M. E. (2015). Social structure, infectious diseases, disasters, secularism, and cultural change in America. Psychological Science, 26(3), 311–324. pmid:25656275

41 Yuval Noah Harari, (Sapiens) A Brief History of Humankind, London 2014, ISBN 978-1846558238

42 Lider.hr, https://lider.media/poslovna-scena/svijet/infografika-sve-pandemije-kroz-povijest-130435 [accessed 23.10.2021]

43 Potter CW (October 2001). “A history of influenza”. Journal of Applied Microbiology. 91 (4): 572–579. doi:10.1046/j.1365-2672.2001.01492.x. PMID 11576290.

44 CDC, https://www.cdc.gov/flu/symptoms/flu-vs-covid19.htm [accessed 23.09.2021]

45 Vijgen L, Keyaerts E, Moes E, Thoelen I, Wollants E, Lemey Ph, Vandamme A, and Van Ranst M (2005). Complete Genomic Sequence of Human Coronavirus OC43: Molecular Clock Analysis Suggests a Relatively Recent Zoonotic Coronavirus Transmission Event, Journal of Virology, 2005, p. 1595–1604 Vol. 79, No. 3, doi:10.1128/JVI.79.3.1595–1604.2005

46 Crookshank, E. M. 1897. Infectious pleuro-pneumonia, p. 239-248. In E. M. Crookshank (ed.), A textbook of bacteriology including the etiology and prevention of infective diseases. W. B. Saunders, Philadelphia.

47 Storz, J., L. Stine, A. Liem, and G. A. Anderson.1996. Coronavirus isolation from nasal swab samples of cattle with signs of respiratory tract disease after shipping. J. Am. Vet. Med. Assoc. 208:1452–1456.

48 Anonymous. 1958. Influenza 1889 and 1957. Lanceti:833-835, cited in: Vijgen et al. (2005)

49 Sisley, R. 1891. The epidemic of 1889-1890. Bokhara. St. Petersburgh. Berlin, p. 47–53. In R. Sisley (ed.), Epidemic influenza: notes on its origin and method of spread. Longmans, Green, and Co., London, United Kingdom.

50 Mulder, J., and N. Masurel. 1958. Pre-epidemic antibody against 1957 strain of Asiatic influenza in serum of older people living in The Netherlands. Lanceti: 810–814.

51 Arbour, N., R. Day, J. Newcombe, and P. J. Talbot. 2000. Neuroinvasion by human respiratory coronaviruses. J. Virol.74:8913–8921.

52 Valleron, A.J., Cori, A., Valtat, S., Meurisse, S., Carrat, F., and Boëlle, P.Y. (2010) Transmissibility and geographic spread of the 1889 influenza pandemic. Proc Natl Acad Sci USA 107: 8778–8781.

53 Dr. Parsons, Report on the Influenz Epidemic of 1889-90 - Great Britain, Local Government Board, Henry Franklin Parsons – Google Books).

54 Kousoulis, Antonis A; Tsoucalas, Gregory; (2017) Infection, contagion and causality in Colonial Britain: the 1889-90 influenza pandemic and the British Medical Journal. Le infezioni in medicina, 25 (3). pp. 285-291. ISSN 1124-9390

55 1911 Encyclopædia Britannica/Influenza – Wikisource

56 Leyden and Guttmann, 1892 https://collections.nlm.nih.gov/catalog/nlm:nlmuid-64820270R-bk

57 Bénézit, F., Le Turnier, P., Declerck, C., Paillé, C., Revest, M., Dubée, V., et al. (2020) Utility of hyposmia and hypogeusia for the diagnosis of COVID-19. Lancet Infect Dis 20: 1014–1015.

58 Telenti, A., Arvin, A., Corey, L., Corti, D., Diamond, M.S., García-Sastre, A., et al. (2021) After the pandemic: perspectives on the future trajectory of COVID-19. Nature 596: 495–504. https://doi.org/10.1038/s41586-021-03792-w

59 Rozen, T.D. (2020) Daily persistent headache after a viral illness during a worldwide pandemic may not be a new occurrence: Lessons from the 1890 Russian/Asiatic flu. Cephalalgia 40: 1406–1409.

60 Laura Spinney, The Spanish Flu of 1918 and How It Changed the World; Adam Kucharski, The Rules of Contagion: Why Things Spread--And Why They Stop.

61 Martin C J Bootsma et al., The effect of public health measures on the 1918 influenza pandemic in U.S. cities 2007, DOI: 10.1073/pnas.0611071104

62 Brian G., COVID-19 Update: Knowledge Is Power, But Compassion Is Lacking, https://www.myeloma.org/blog/covid-19-update-knowledge-power-compassion-lacking [accessed 02.01.2022]

63 Opca i nacionalna enciklopedija, Zagreb 2006.

64 Gooding P. (2012). Mass digitization and the garbage dump: The conflicting needs of quantitative and qualitative methods. Literary and Linguistic Computing, 28(3), 425–431; See also

65 Pettit M. (2016). Historical time in the age of big data: Cultural psychology, historical change, and the Google Books Ngram Viewer. History of Psychology, 19(2), 141. pmid:27100927

66 Pechenick, Eitan Adam; Danforth, Christopher M.,, Dodds, Peter Sheridan; Barrat, Alain (7 October 2015). “Characterizing the Google Books Corpus: Strong Limits to Inferences of Socio-Cultural and Linguistic Evolution”. PLOS ONE. 10 (10): e0137041. doi:10.1371/journal.pone.0137041.

67 Zhang, Sarah. “The Pitfalls of Using Google Ngram to Study Language”. WIRED. (2017-05-24.)

68 Hilpert, Martin and Stefan Gries. 2009. Assessing Frequency Changes in multistage Diachronic Corpora: Applications for Historical Corpus Linguistics and the Study of Language Acquisition. Literary and Linguistic Computing, 24(4), 385–401. DOI: 10.1093/llc/fqn012

69 Danah Boyd & Kate Crawford (2012) CRITICAL QUESTIONS FOR BIG DATA, Information, Communication & Society, 15:5, 662–679, DOI: 10.1080/1369118X.2012.678878

70 Davis, Mark. 2014. Making Google Books n-grams Useful for a Wide Range of Research on Language Change. International Journal of Corpus Linguistics 19(3), 401–16.

71 Lakoff, Robin. 2013. What Words Don’t Tell Us., http://blogs.berkeley.edu/author/rlakoff/ [accessed 23.12.2021]

72 Solovyev V.D., Bochkarev V.V., Akhtyamova S.S. (2020) Google Books Ngram: Problems of Representativeness and Data Reliability. In: Elizarov A., Novikov B., Stupnikov S. (eds) Data Analytics and Management in Data Intensive Domains. Communications in Computer and Information Science, vol 1223. Springer, Cham. https://doi.org/10.1007/978-3-030-51913-1_10

